# Proteomic Profiling of Nasal Fluid from the Brain-Nose Interface using Mass Spectrometry

**DOI:** 10.64898/2026.07.20.26358433

**Authors:** Stephan A. Müller, Xiao Feng, Maria Babu, Sabine Mertzig, Richard Metzler, Hilary Wunderlich, Gunter Weiss, Mohammad Bashiri, Oliver Peters, Lutz Frölich, Jens Wiltfang, Timo J. Oberstein, Marion San Nicoló, Stefan F. Lichtenthaler, Mareike Albert

## Abstract

Nasal fluid collected at the brain-nose interface (BNI) may provide a minimally invasive window into central nervous system (CNS) biology, but its suitability for deep, reproducible proteomics remains unclear. Here, we establish and benchmark a standardized liquid chromatography-tandem mass spectrometry workflow for BNI-proximal nasal fluid and compare its proteome to matched cerebrospinal fluid (CSF) from 40 individuals with cognitive impairment. Using pooled nasal fluid, we identified single-pot, solid-phase sample preparation combined with data-independent acquisition (DIA) as the optimal strategy, with low coefficients of variation. In cognitively impaired cohort, DIA-based proteomics on a timsTOF pro quantified on average 5,210 proteins in nasal fluid and 1,800 in CSF, with 4,901 nasal fluid proteins detected in at least 75% of samples. Nasal fluid contained nearly 50% of the CSF proteome (876 shared proteins), including numerous CNS- and neurodegeneration-associated species. The result of this exploratory study position BNI-derived nasal fluid as a robust, proteome-rich biofluid for proteomics-driven CNS biomarker research.

## Introduction

Biomarker discovery in neurodegenerative disease relies on proteomic technologies capable of characterizing complex biological specimens. Proteomics has been central to defining molecular signatures linked to disease mechanisms, progression, and patient stratification, but these efforts are constrained by the need for biofluids that are both mechanistically informative and suitable for large-scale or longitudinal sampling. Cerebrospinal fluid (CSF) is the reference biofluid for assessing central nervous system (CNS) pathology, including established Alzheimer’s disease (AD) biomarkers such as amyloid-β and tau^1,2^. However, lumbar puncture limits routine use, repeated measurements, and population-based studies, motivating the search for alternative, non-invasively collected specimens which are compatible with robust, reproducible mass spectrometry-based proteomics^3^.

Nasal fluid (NF) collected at the brain-nose interface (BNI) represents a promising yet underexplored candidate biofluid for CNS biomarker discovery. The BNI comprises three main structures: the cribriform plate as the perforated bony barrier between cranial and nasal cavities, the olfactory bulb above it, and the olfactory mucosa of the upper nasal cavity below it (Fig. 1A)^4–6^. Olfactory receptor neurons project from the nasal mucosa through the cribriform plate to the olfactory bulb, creating a direct anatomical continuum between nasal mucosa and CNS through which CNS-derived proteins and CSF can reach the olfactory mucosa and nasal mucosal lymphatics^5,7–9^. Consistent with this connection, AD biomarkers have been reported to be elevated in nasal samples from AD patients compared with healthy controls^7–9^.

**Figure 1.**
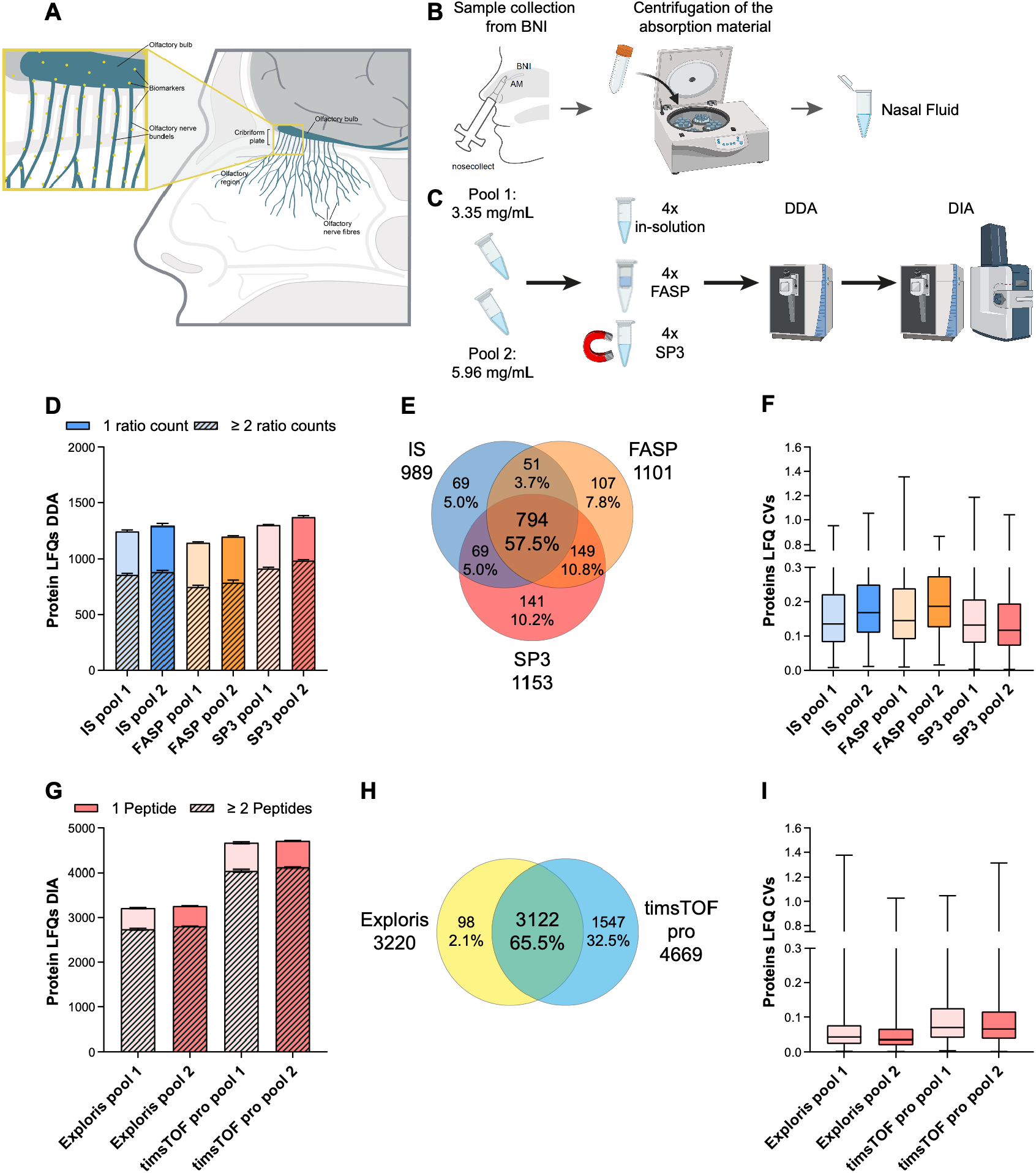
Collection of nasal fluid (NF) samples and method optimization for mass spectrometry-based proteomics. **A)** Schematic representation of the brain-nose interface (BNI), including the olfactory bulb, cribriform plate, and olfactory epithelium in the superior nasal cavity, illustrating the anatomical continuum between the CNS and the nasal mucosa. *(Reproduced from San Nicoló, Marion, et al. “Novel, standardized sample collec2on from the brain-nose interface*.*” Methods 234 (2025): 233-241*.,^*6*^ *under the terms of the Creative Commons Attribution 4*.*0 Interna2onal License (CC BY 4*.*0) (http://creativecommons.org/licenses/by/4.0/))* **B**) Workflow of NF collection at the BNI using the nosecollect® device. The absorbent material is positioned in the upper nasal cavity adjacent to the BNI for a defined sampling period, then retrieved and transferred into collection tubes. NF is subsequently recovered from the absorbent material by centrifugation, followed by downstream processing and mass spectrometry-based proteomic analysis. (*Adapted in part from San Nicoló, Marion, et al. “Novel, standardized sample collec2on from the brain-nose interface*.*” Methods 234 (2025): 233-241*.,*20 under the terms of the Creative Commons Attribution 4*.*0 Interna2onal License (CC BY 4*.*0) (http://creativecommons.org/licenses/by/4.0/)) (Created with BioRender (https://BioRender.com))* **C**) Two NF pools with low and high protein concentrations were utilized for method optimization in quadruplicates comparing in-solution digestion, filter aided sample preparation (FASP) and single-pot, solid-phase, sample-preparation (SP3) with DDA measurements on an Exploris 480 mass spectrometer. The samples of the best performing preparation method were further analyzed by DIA on an Exploris 480 and a timsTOF pro mass spectrometer *(Created with BioRender (https://BioRender.com))*. **D**) The numbers of protein quantifications with one and two ratio counts show the highest values for in-solution (IS) and SP3 digestion. **E**) Overlap of protein quantifications in at least three out of four replicates for both high and low concentration pools for all sample preparation methods. **F**) The coefficient of variation (CV) shows the lowest values for the SP3 method. **G**) Comparison of protein quantifications with one and two peptides using DIA on an Exploris 480 and a timsTOF pro mass spectrometer. **H**) Venn diagram of proteins quantified with DIA in 75% of all samples. **I**) A box plot (min to max) of CV values shows good reproducibility of protein quantification with DIA for both instruments.

Within the BNI, CNS-related proteins are likely less affected by systemic dilution and metabolism than in peripheral blood, making BNI-proximal NF an attractive matrix for proteomics-based biomarker discovery^10,11^. In previous studies, nasal proteomics has largely focused on rhinological cohorts and has not specifically targeted the BNI, limiting the systematic assessment of the CNS-linked proteome in NF ^12–16^. To access BNI in a targeted manner, San Nicoló et al. developed a dedicated NF collection device (nosecollect®) for targeted, minimally invasive, standardized sampling at BNI, suitable for repeated and large-scale studies with reduced pre-analytical variability^6^. In this technical brief, sample preparation and liquid chromatography-tandem mass spectrometry (LC-MS/MS)-based analysis of nosecollect®-derived NF from the BNI are optimized, and then applied to 40 NF and matched CSF samples from individuals with cognitive impairment, to evaluate the technical feasibility and potential utility of NF as a minimally invasive specimen for proteomics-driven CNS biomarker research. We show that BNI-proximal NF measured with optimized mass spectrometry strategy yields a CSF-overlapping, neurodegeneration-enriched proteome, establishing NF as a discovery-stage biofluid for CNS biomarker research.

## Results and Discussion

NF collected at the BNI with nosecollect® provides an easily accessible biofluid with the potential to identify protein biomarkers (Fig. 1A, B). To optimize and generate an in-depth proteomics analysis of NF, we first tested different sample preparation methods using technical quadruplicates of two healthy NF pools with low and high total protein concentrations, respectively, using data-dependent acquisition (DDA) on a Vanquish Neo Orbitrap Exploris 480 system (Fig. 1C). In-solution and single-pot, solid-phase sample preparation (SP3) provided similar protein ID numbers, while filter-aided sample preparation (FASP) yielded the lowest (Fig. 1D). Taking only proteins into account which were identified in 3 out of 4 replicates in both NF pools, SP3 showed the best performance with 1153 proteins detected (Fig. 1E). Additionally, SP3 provided the best quantification reproducibility with the lowest coefficient of variation (CV) values (Fig. 1F). Next, we used the SP3 digested NF samples to evaluate the gain of identifications using data-independent acquisition (DIA) on an Exploris 480 and a timsTOF pro mass spectrometer (Fig. 1C). The application of DIA more than doubled the ID numbers on the Exploris 480 compared to DDA, while the timsTOF pro showed the better performance with 45% more IDs on average (Fig 1G) and increased the number of proteins quantified in 75% of all samples (Fig. 1H) However, DIA on the Exploris 480 had the better quantification reproducibility, while results on the timsTOF pro also displayed very good CV values (Fig. 1I).

Next, we aimed to compare the proteome of NF and CSF from the 40 individuals with cognitive impairment (age mean ± standard deviation (SD), (median, min-max): 70.5 ± 9.3, (73.0, 49.0-86.0); male: 21 (52.5%); Mini-Mental Status Test mean ± SD, (median, min-max): 25.4 ± 4.1, (27.0, 10.0-30.0). We applied DIA on the timsTOF Pro mass spectrometer as the more sensitive method. On average, 1800 and 5210 proteins were quantified in CSF and NF with at least one peptide, respectively (Fig. 2A). The number of peptides and proteins detected across each individual sample clearly demonstrates the consistently higher proteome content in NF compared to CSF (SI Fig. 1A-D). When filtering for proteins quantified in at least 75% of all samples, we observed an overlap of 876 proteins while 619 and 4025 proteins were exclusively quantified in CSF and NF, respectively (Fig. 2B). CSF proteomics mainly quantified secreted and proteolytically shed membrane proteins, while NF proteomics showed a higher percentage of proteins annotated for cytoplasm and nucleus (Fig. 2C), which might indicate a stronger cellular turnover at the BNI as well as the residual mucosal cells in the samples. Differences in subcellular location are also displayed in a gene ontology enrichment analysis for cellular component compared to the whole human proteome as a background (Fig. 2D). NF showed a stronger enrichment for cytoplasm and nucleus indicating a higher cellular content, whereas CSF was more enriched for membrane and cell surface proteins. tiet, both showed significant enrichments for extracellular exosome, region and space. For biological function, NF and CSF showed the highest enrichment for protein transport and cell adhesion, respectively, while both were significantly enriched for immune system process (Fig. 2E). Next, we checked the correlation of paired NF and CSF label-free quantification (LFQ) intensities from the different individuals and discovered 92 and 8 proteins with a positive and negative correlation (p < 0.05), respectively (Fig. 2F, G). Among the proteins with positive correlation, 44 proteins were associated with immune response including mainly immunoglobulin, but also CCL14, APP, CFHR2 and 5, MST1, and major histocompatibility class I proteins HLA-A and HLA-C. We also identified 68 brain elevated proteins according to the human protein atlas among the shared proteome (Fig. 2F), which might be derived from brain tissue.

**Figure 2.**
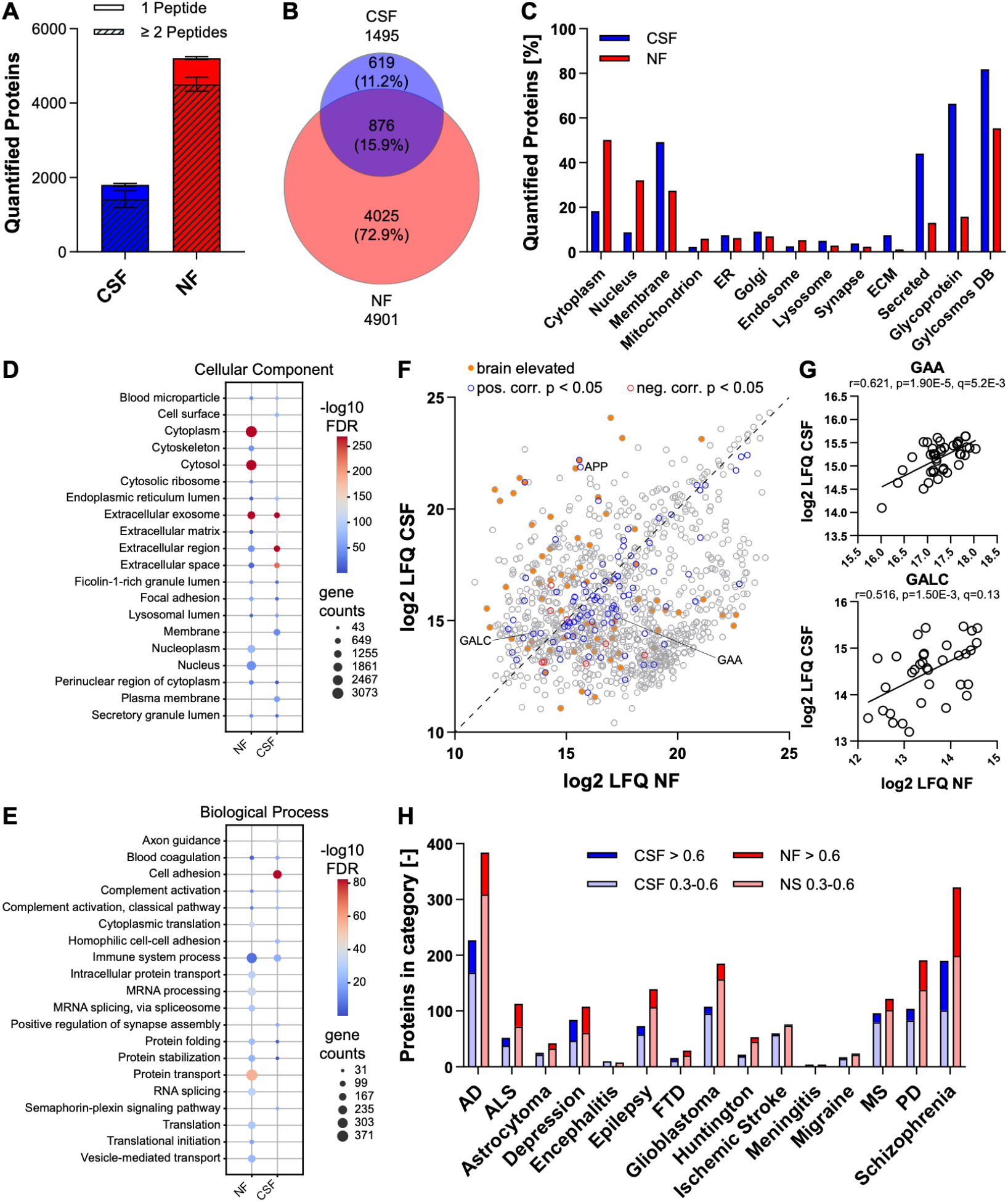
Proteomic comparison of CSF and nasal fluid (NF) from 40 cognitively impaired individuals using the optimized mass spectrometry method. **A**) Number of proteins identified by at least two or one peptide in CSF and NF. **B**) Venn diagram of proteins quantified in at least 75% of all samples in CSF and NF. **C**) Bar graph of UniProt subcellular locations as well as UniProt keyword glycoprotein and the Glycosmos DB annotation for proteins quantified in 75% of all samples. Bubble plots of the gene ontology enrichment analysis for proteins quantified in 75% of the samples according to cellular component (**D**) and biological process (**E**) with the webtool DAVID (CC_direct and BP_direct). **F**) Scaver plot of average log2 LFQ intensities of NF and CSF. Proteins with a positive or negative correlation with a p-value less than 0.05 are indicated as blue and red dots, respectively. Proteins elevated in brain tissue according to the human protein atlas are indicated in orange. **G**) Scaver plots of log2 LFQ intensities of the proteins GAA and GALC for each donor. **H**) Bar plot of proteins associated with neurological diseases (AD: Alzheimer’s disease, FTD: frontotemporal dementia, MS: multiple sclerosis, PD: Parkinson’s disease) according to the DISGENET database with a score of 0.3-0.6 (low to medium confidence association) or more than 0.6 (high confidence association).

Furthermore, to explore proteins relevant in neurological disorders, we used the DISGENET database for gene disease association^17,18^ (Fig. 2H). These proteins represent potential targets for CNS biomarker and basic research. Both body fluids include several hundreds of proteins related to different diseases offering a potential to obtain relevant disease related information or even to identify biomarkers using state-of-the-art proteomics analyses.

NF collected at the BNI with the standardized nosecollect® system emerges from this work as a technically feasible and biologically rich matrix to investigate neurological diseases. Our data demonstrates that NF proteomics is a robust and reproducible technique with 81-91% of detected proteins showing CVs below 15% using NF pools. The data completeness was also excellent for the proteomic comparison of paired NF-CSF samples, with 4901 proteins in 75% of all NF samples. More sensitive mass spectrometers offering a larger dynamic range such as the timsTOF ultra and Orbitrap Astral will even provide significantly more detailed analyses with greater depth. A central finding is the substantial compositional overlap between NF and CSF. More than half of the CSF proteome was detectable in NF with 876 shared proteins, out of which 92 showing a positive correlation of LFQ intensities. Functional enrichment analysis revealed strong representation of CNS-relevant categories for both body fluids, including proteins annotated to Alzheimer’s disease, Parkinson’s disease, and broader neurodegenerative disease pathways (Fig. 2D, E, H). These findings support a functional CNS-nasal connection and align with anatomical models of CSF drainage along olfactory routes^5^, while the incomplete overlap between CSF and NF proteomes suggests additional selective transport, dilution, or local processing.

Our findings extend previous mass spectrometry-based studies of nasal mucus and nasal lavage in predominantly rhinological cohorts, which typically achieved more limited proteome depth, ranging from ~20-430 identified proteins using 2DE/MALDI-TOF, early ion-trap LC-MS/MS or nano-LC-MS/MS platorms^12–16^ to ~953 proteins with TMT-based quantitation^19^ and up to ~2,000 proteins in a recent high-resolution DDA study^20^- and largely reported mucosal barrier proteins, and innate immune and inflammatory mediators. A deep shotgun LC-MS/MS analysis of olfactory cle| mucus by Yoshikawa et al. cataloged ~3,000 proteins but was limited to healthy individual samples^21^.Moreover, none of these previous studies integrated CSF analysis, did not focus on systematic characterization of CNS/neurodegeneration-linked proteins in NF or interrogate the samples targetedly collected from BNI with nosecollect®. By contrast to the past studies, our dataset reveals a greater depth of coverage in NF (5210 proteins on average, 4901 proteins in 75% of all NF samples), significant overlap with CSF proteome and a broader, systematically quantified panel of neurodegeneration-associated species in both NF and CSF, suggesting that BNI-proximal NF collected with nosecollect® captures CNS-linked proteomic information comprehensively. Whether the shared and disease-associated proteins in NF are truly CNS-derived, reflect secondary CSF transfer, or arise from local or systemic sources remains unresolved.

Nevertheless, also limitations of this work should be considered when interpreting these findings: The cohort was modest in size (n = 40 matched NF-CSF pairs). The proteomic dynamic range was another constraint in both matrices: despite deep coverage, low-abundance canonical AD biomarkers (NEFL, MAPT) were not reliably detected by untargeted LC-MS/MS in either fluid. This shortcoming might be solved with a more sensitive MS setup or with targeted proteomics techniques.

Despite these limitations, our data shows that BNI-derived NF has the potential to identify biomarkers for CNS biomarker research in addition to CSF. We establish an optimal LC-MS/MS workflow suitable for NF allowing detection with high proteome depth. More than half of the CSF proteome is detectable in NF, with hundreds of shared proteins, including numerous potentially brain-derived and neurodegeneration-related species, jointly observed across fluids. Together, these findings highlight NF as an easily accessible and patient-friendly body fluid with strong potential for generating high-quality biological data and enabling biomarker discovery in large cohort studies of neurological disorders

## Methods

A detailed version of the material and methods section can be found in the supplementary material. Participant recruitment and sampling were performed within a clinically approved study protocol authorized by Ethik-Kommission der Bayerischen Landesärztekammer (approval no. 21112) and registered on ClinicalTrials.gov (identifier NCT05791552). NF samples from the BNI region was collected using the nosecollect® device, a dedicated nasal sampling tool targeting fluid collection from the upper nasal cavity^6^. CSF was obtained by lumbar puncture according to standard clinical procedures.

For method optimization, two pools of healthy NF samples with high (5.96 mg/mL) and low (3.35 mg/mL) total protein concentration were used. Four technical replicates of 20 µg per sample were prepared with trypsin and LysC using in-solution (IS) digestion with 0.1% (w/v) sodium deoxycholate^22^, FASP^23^ and SP3^24^. Protein digestion was performed with 125 ng LysC and 250 ng trypsin (Promega). The best sample preparation method for NF was identified using a Top 30 DDA method on a Vanquish Neo - Orbitrap Exploris 480 LC-MS/MS system. A|erwards, samples from SP3 digestion were analyzed using DIA on a Vanquish Neo - Orbitrap Exploris 480 LC-MS/MS system and a nanoElute timsTOF pro system.

For matched NF-CSF comparison using cognitively impaired patient samples, a volume of 15 µL of CSF per sample was transferred to a 96-well plate (AB2800 SuperPlate, ThermoFisher Scientific). For NF samples, 20 µg per sample were transferred to a 96-well plate and H_2_O was added to a final volume of 15 µL per well. Sample preparation was performed using the automated SP3 protocol^25^ using LysC trypsin double digestion on a Bravo pipetting robot (Agilent, US) equipped with a heater, a shaker and a Magnum FLX 96-well magnetic plate (Alpaqua, US, SKU: A000400) with the method SP3_1.01 provided by Agilent. Samples were analyzed using a diaPASEF method on a nanoElute timsTOF pro system.

The DDA data was analyzed with the so|ware Maxquant Version 2.5.1.0^26^ using a one protein per gene database from Homo sapiens (UniProt, 20662 entries, release date: 2024-07-24) including the Maxquant contamination database with default settings for label-free quantification (LFQ). The DIA raw data was analyzed with DIA-NN versions 1.8.1 (method test) and 1.9.2 (NF, CSF comparison)^27^. The raw data was searched against a one protein per gene database from Homo sapiens (UniProt, 20662 entries, release date: 2024-07-24) and a database with potential contaminants (123 entries) using a library free search.

The so|ware Perseus version 2.1.3.0^28^ was used for further data analysis. Enrichment analyses were performed with DAVID webtool. Data visualization was performed with python using the Matplotlib package or with Graphpad Prism version 10.6.1. Gene disease associations from DISGENET database (https://www.disgenet.com) were downloaded at 2026-01-20^17,18^.

## Abbreviations

AD: Alzheimer’s disease
BNI: brain-nose interface
CNS: central nervous system
CSF: cerebrospinal fluid
CV: coefficient of variation
DDA: Data-dependent acquisition
DIA: Data-independent acquisition
FASP: Filter-aided sample preparation
IS: In-solution
LC-MS/MS: liquid chromatography-tandem mass spectrometry
LFQ: label-free quantification
NF: nasal fluid
SP3: Single-pot, solid-phase sample preparation
SD: Standard deviation

## Declarations

## Acknowledgements

Biomed advisor AI was used for improving language. BioRender was used for creation of images (https://BioRender.com).

## Author contributions

SAM: methodology, investigation, data analysis, visualization, original dra| writing and review; XF: data analysis, review; MB: original dra| writing and review; SM: methodology, review; RM: data analysis, review; HW: methodology; GW: methodology; MoB: review; OP: resources; LF: resources; JW: resources; TJO: resources; MSN: methodology; SFL: conceptualization, methodology, review, supervision; MA: conceptualization, methodology, review, supervision

## Data availability

Due to ongoing patent applications and institutional restrictions, the underlying datasets in this study cannot be made available.

## References

1. Jack CR, Andrews JS, Beach TG, et al. Revised criteria for diagnosis and staging of Alzheimer’s disease: Alzheimer’s Association Workgroup. Alzheimers Dement. 2024;20(8):5143–5169. doi:10.1002/alz.13859

2. Jack CR, Bennev DA, Blennow K, et al. NIA-AA Research Framework: Toward a biological definition of Alzheimer’s disease. Alzheimers Dement. 2018;14(4):535–562. doi:10.1016/j.jalz.2018.02.018

3. Jung DH, Son G, Kwon OH, Chang KA, Moon C. Non-Invasive Nasal Discharge Fluid and Other Body Fluid Biomarkers in Alzheimer’s Disease. Pharmaceu2cs. 2022;14(8):1532. doi:10.3390/pharmaceutics14081532

4. Johnston M, Zakharov A, Papaiconomou C, Salmasi G, Armstrong D. Evidence of connections between cerebrospinal fluid and nasal lymphatic vessels in humans, non-human primates and other mammalian species. Cerebrospinal Fluid Res. 2004;1:2. doi:10.1186/1743-8454-1-2

5. Mehta NH, Sherbansky J, Kamer AR, et al. The Brain-Nose Interface: A Potential Cerebrospinal Fluid Clearance Site in Humans. Front Physiol. 2022;12:769948. doi:10.3389/fphys.2021.769948

6. San Nicoló M, Mertzig S, Berghaus A, et al. Novel, standardized sample collection from the brain-nose interface. Methods. 2025;234:233–241. doi:10.1016/j.ymeth.2024.12.012

7. Yoo SJ, Son G, Bae J, et al. Longitudinal profiling of oligomeric Aβ in human nasal discharge reflecting cognitive decline in probable Alzheimer’s disease. Sci Rep. 2020;10(1):11234. doi:10.1038/s41598-020-68148-2

8. Kim YH, Lee SM, Cho S, et al. Amyloid beta in nasal secretions may be a potential biomarker of Alzheimer’s disease. Sci Rep. 2019;9(1):4966. doi:10.1038/s41598-019-41429-1

9. Passali GC, Politi L, Crisanti A, Loglisci M, Anzivino R, Passali D. Tau Protein Detection in Anosmic Alzheimer’s Disease Patient’s Nasal Secretions. Chemosens Percept. 2015;8(4):201–206. doi:10.1007/s12078-015-9198-3

10. Ozaki T, Yoshino Y, Tachibana A, et al. Metabolomic alterations in the blood plasma of older adults with mild cognitive impairment and Alzheimer’s disease (from the Nakayama Study). Sci Rep. 2022;12(1):15205. doi:10.1038/s41598-022-19670-y

11. Huang S, Wang YJ, Guo J. Biofluid Biomarkers of Alzheimer’s Disease: Progress, Problems, and Perspectives. Neurosci Bull. 2022;38(6):677–691. doi:10.1007/s12264-022-00836-7

12. Ghafouri B, Irander K, Lindbom J, Tagesson C, Lindahl M. Comparative proteomics of nasal fluid in seasonal allergic rhinitis. J Proteome Res. 2006;5(2):330–338. doi:10.1021/pr050341h

13. Tewfik MA, Laverich M, DiFalco MR, Samaha M. Proteomics of nasal mucus in chronic rhinosinusitis. Am J Rhinol. 2007;21(6):680–685. doi:10.2500/ajr.2007.21.3103

14. Tomazic PV, Birner-Gruenberger R, Leitner A, Obrist B, Spoerk S, Lang-Loidolt D. Nasal mucus proteomic changes reflect altered immune responses and epithelial permeability in patients with allergic rhinitis. J Allergy Clin Immunol. 2014;133(3):741–750. doi:10.1016/j.jaci.2013.09.040

15. Benson LM, Mason CJ, Friedman O, Kita H, Bergen HR, Plager DA. Extensive fractionation and identification of proteins within nasal lavage fluids from allergic rhinitis and asthmatic chronic rhinosinusitis patients. J Sep Sci. 2009;32(1):44–56. doi:10.1002/jssc.200800474

16. Tomazic PV, Birner-Gruenberger R, Leitner A, Spoerk S, Lang-Loidolt D. Seasonal proteome changes of nasal mucus reflect perennial inflammatory response and reduced defence mechanisms and plasticity in allergic rhinitis. J Proteomics. 2016;133:153–160. doi:10.1016/j.jprot.2015.12.021

17. Piñero J, Ramírez-Anguita JM, Saüch-Pitarch J, et al. The DisGeNET knowledge platorm for disease genomics: 2019 update. Nucleic Acids Res. 2020;48(D1):D845–D855. doi:10.1093/nar/gkz1021

18. Piñero J, Corvi J, Rykova N, et al. DISGENET: Accelerating Data-Driven Discovery in Disease Genomics and Therapeutic Development. Preprint posted online January 5, 2026. doi:10.64898/2026.01.05.697749

19. Wang H, GoÖries J, Barrenäs F, Benson M. Identification of novel biomarkers in seasonal allergic rhinitis by combining proteomic, multivariate and pathway analysis. PLoS One. 2011;6(8):e23563. doi:10.1371/journal.pone.0023563

20. Kim YS, Han D, Kim J, et al. In-Depth, Proteomic Analysis of Nasal Secretions from Patients With Chronic Rhinosinusitis and Nasal Polyps. Allergy Asthma Immunol Res. 2019;11(5):691–708. doi:10.4168/aair.2019.11.5.691

21. Yoshikawa K, Wang H, Jaen C, et al. The human olfactory cle| mucus proteome and its age-related changes. Sci Rep. 2018;8(1):17170. doi:10.1038/s41598-018-35102-2

22. Pigoni M, Wanngren J, Kuhn PH, et al. Seizure protein 6 and its homolog seizure 6-like protein are physiological substrates of BACE1 in neurons. Mol Neurodegener. 2016;11(1):67. doi:10.1186/s13024-016-0134-z

23. Wiśniewski JR, Zougman A, Nagaraj N, Mann M. Universal sample preparation method for proteome analysis. Nat Methods. 2009;6(5):359–362. doi:10.1038/nmeth.1322

24. Hughes CS, Moggridge S, Müller T, Sorensen PH, Morin GB, Krijgsveld J. Single-pot, solid-phase-enhanced sample preparation for proteomics experiments. Nat Protoc. 2019;14(1):68–85. doi:10.1038/s41596-018-0082-x

25. Müller T, Kalxdorf M, Longuespée R, Kazdal DN, Stenzinger A, Krijgsveld J. Automated sample preparation with SP3 for low-input clinical proteomics. Mol Syst Biol. 2020;16(1):e9111. doi:10.15252/msb.20199111

26. Cox J, Hein MY, Luber CA, Paron I, Nagaraj N, Mann M. Accurate proteome-wide label-free quantification by delayed normalization and maximal peptide ratio extraction, termed MaxLFQ. Mol Cell Proteomics. 2014;13(9):2513–2526. doi:10.1074/mcp.M113.031591

27. Demichev V, Messner CB, Vernardis SI, Lilley KS, Ralser M. DIA-NN: neural networks and interference correction enable deep proteome coverage in high throughput. Nat Methods. 2020;17(1):41–44. doi:10.1038/s41592-019-0638-x

28. Tyanova S, Temu T, Sinitcyn P, et al. The Perseus computational platorm for comprehensive analysis of (prote)omics data. Nat Methods. 2016;13(9):731–740. doi:10.1038/nmeth.3901

