## Supplementary figure and methods for "Proteomic Profiling of Nasal Fluid from the Brain-Nose Interface using Mass Spectrometry"

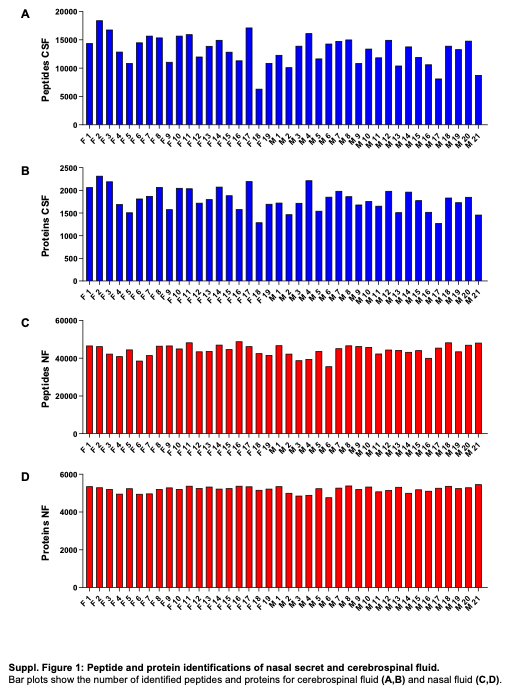

**Supplementary Figure 1: Peptide and protein identifications of nasal fluid (NF) and cerebrospinal fluid (CSF).** Bar plots show the number of identified peptides and proteins for cerebrospinal fluid (A,B) and nasal fluid (C,D)

### Materials and Methods

**Participant recruitment**

The research was conducted under the ethical and legal framework of the Declaration of Helsinki. Participant recruitment and sampling were performed within a clinically approved study protocol authorized by Ethik-Kommission der Bayerischen Landesärztekammer (approval no. 21112) and registered on ClinicalTrials.gov (identifier NCT05791552). Because lumbar puncture in neurologically healthy volunteers is ethically difficult to justify in the absence of clinical indication, the study population for nasal fluid (NF)- cerebrospinal fluid (CSF) matched study was restricted to individuals with cognitive impairment, in whom CSF collection forms part of the routine diagnostic evaluation. CSF specimens and NF were obtained from patients with cognitive deficits at collaborating memory clinics. For method optimisation, pooled NF samples from healthy subjects were utilised. Written informed consent was obtained from all participants before any study-related procedures.

**Sample collection**

NF from the brain-nose interface (BNI) region was collected using the nosecollect® device, a dedicated nasal sampling tool targeting fluid collection from the upper nasal cavity. As the device is inserted through the nostril, the absorbent material that absorbs the nasal fluid remains enclosed within the distal tip of the device. As the tip of the device approaches the vicinity of BNI, absorbent material is deployed in a minimally invasive manner. The absorbent material is retrieved from the BNI after incubation by slowly pulling out the thread attached to it which extends out through the nostril. The technical performance and applicability of nosecollect® for BNI-targeted sampling have been described by San Nicoló et al^1^.

Collections were performed in a clinical setting by staff trained in nosecollect® handling. After sampling, absorbent materials from both nasal cavities were pooled into Eppendorf Protein LoBind tubes. CSF was obtained by lumbar puncture according to standard clinical procedures. Absorbent materials with nasal fluid and CSF were frozen on site at −80 °C and shipped on dry ice to biobank facility, where NF was eluted from the matrices and all samples were stored at −80 °C. For downstream protein profiling by mass spectrometry, aliquots of NF and CSF were shipped on dry ice from the biobank to measurement site and stored at −80 °C until analysis.

**Method optimization**

For method optimization, two pools of healthy NF samples with high (5.96 mg/mL) and low (3.35 mg/mL) total protein concentration were used. Four technical replicates of 20 µg per sample were prepared with trypsin and LysC using in-solution (IS) digestion with 0.1% (w/v) sodium deoxycholate^2^, filter aided sample preparation (FASP)^3^ and single-pot, solid-phase, sample-preparation (SP3)^4^. Protein digestion was performed with 125 ng LysC and 250 ng trypsin (Promega).

To identify the best sample preparation method for NF, samples were measured on a Vanquish Neo - Orbitrap Exploris 480 LC-MS/MS system using a Top 30 data dependent acquisition (DDA) method. 400 ng were separated on a two column setup (Thermo C18 Trap cartridge 300 µm × 0.5 cm and PepSep C18 15cm x 75 µm x 1.9 µm) (A) using a binary gradient of water and acetonitrile (B) containing 0.1% formic acid at flow rate of 300 nL/min (0 min, 3.2% B; 0.7 min, 3.6% B; 1 min, 5%; 38 min, 16% B; 56.9 min, 28% B; 57.4 min, 44% B; 57.9 min, 79.2%) and a column temperature of 50°C. MS1 spectra were acquired at a resolution of 60,000 and AGC of 300% covering a m/z range from 300-1400. The top 30 most intense ions with a charge state of 2-5 were chosen for collision induced dissociation (isolation width 2 m/z, normalized collision energy 30%, resolution 15,000, AGC 100%, dynamic exclusion 50 s).

The samples of the best method were measured again using data independent acquisition (DIA) on two different systems, Vanquish Neo - Orbitrap Exploris 480 and a nanoElute - timsTOF pro system, to evaluate the improvement achieved with DIA.

For DIA on the Vanquish Neo - Orbitrap Exploris 480 LC-MS/MS system, using a binary gradient of water and acetonitrile (B) containing 0.1% formic acid at flow rate of 300 nL/min (0 min, 4.8% B; 2 min, 4.8% B; 62 min, 24.8% B; 67 min; 40% B; 72 min; 76% B) and a column temperature of 50°C. A DIA method with variable window sizes covering a m/z range from 350 to 1001 was applied (20 windows: m/z 380, width 60; m/z 426, width 34; m/z 457, width 30; m/z 485, width 28; m/z 511, width 26; m/z 536, width 26; m/z 560, width 24; m/z 583, width 24; m/z 606, width 24; m/z 629, width 24; m/z 653, width 26; m/z 678, width 26; m/z 705, width 30; m/z 735, width 32; m/z 766, width 32; m/z 798, width 34; m/z 833, width 38; m/z 872, width 42; m/z 915, width 46; m/z 969, width 64). MS1 spectra were acquired at a resolution of 120,000 and an AGC of 300%. MS2 spectra were acquired at a resolution of 30,000, an AGC of 3000, first mass at 200 m/z using a stepped normalized collision energy of 27.5±2.5%.

On the nanoElute timsTOF pro system, a peptide amount of 300 ng per sample for NF or 200 ng per sample for CSF was separated on an in-house packed C18 analytical column (15 cm × 75 µm ID, ReproSil-Pur 120 C18-AQ, 1.9 µm, Dr. Maisch GmbH) using a binary gradient of water and acetonitrile (B) containing 0.1% formic acid at flow rate of 300 nL/min (0 min, 2% B; 2 min, 5% B; 62 min, 24% B; 72 min, 35% B; 75 min, 60%; B78 min, 85%) and a column temperature of 50°C. The nanoHPLC was online coupled to a timsTOF pro mass spectrometer (Bruker, Germany) with a CaptiveSpray ion source (Bruker, Germany). A Data Independent Acquisition Parallel Accumulation–Serial Fragmentation (diaPASEF) method was used for spectrum acquisition. Ion accumulation and separation using Trapped Ion Mobility Spectrometry (TIMS) was set to a ramp time of 100 ms. One scan cycle included one TIMS full MS scan with 26 windows with a width of 27 m/z covering a m/z range of 350-1001 m/z. Two windows were recorded per PASEF scan. This resulted in a cycle time of 1.4 s.

**Proteomic analysis of NF and CSF**

A volume of 15 µL of CSF per sample was transferred to a 96-well plate (AB2800 SuperPlate, ThermoFisher Scientific). For NF samples, 20 µg per sample were transferred to a 96-well plate and H_2_O was added to a final volume of 15 µL per well. For NF samples, a nuclease digestion was performed before starting the automated sample preparation to remove DNA which interferes with the automated sample processing. For this purpose, 5 µL of 100 mM MgCl_2_ and 5 µL of 0.5 U/µL nuclease solution (88216, ThermoFisher Scientific) were added and samples were incubated for 30 min at 37°C.

Sample preparation was performed using the automated single tube, solid phase, sample preparation protocol (SP3)^5^ on a Bravo pipetting robot (Agilent, US) equipped with a heater, a shaker and a Magnum FLX 96-well magnetic plate (Alpaqua, US, SKU: A000400) with the method SP3_1.01 provided by Agilent. For CSF protein digestion, 300 ng LysC (Promega, Germany) and 300 ng trypsin (Promega, Germany) were added in 50 µL of 50 mM ammonium bicarbonate solution. For NF protein digestion, 125 ng LysC and 250 ng trypsin were used. Samples were analyzed with the DIA method mentioned above on a nanoElute timsTOF pro system.

**Data analysis**

The DDA data was analyzed with the software Maxquant Version 2.5.1.0^6^ using a one protein per gene database from Homo sapiens (UniProt, 20662 entries, release date: 2024-07-24) including the Maxquant contamination database. Trypsin was defined as protease. Two missed cleavages were allowed. Oxidation of methionines and acetylation of protein N-termini were defined as variable modifications, whereas carbamidomethylation of cysteines was defined as fixed modification. The peptide mass accuracy was set to 20 ppm for the first search and 20 ppm for the main search. MS/MS spectra were analyzed with a mass accuracy of 20 ppm. Protein and peptide FDR were set to 1%. The software DIA-NN versions 1.8.1 (method test) and 1.9.2 (NF, CSF comparison) were used for label-free quantification (LFQ) of DIA raw data^7^ . The raw data was searched against a one protein per gene database from Homo sapiens (UniProt, 20662 entries, release date: 2024-07-24) and a database with potential contaminants (123 entries) using a library free search. Trypsin was defined as protease and two missed cleavages were allowed. Oxidation of methionines and acetylation of protein N-termini were defined as variable modifications, whereas carbamidomethylation of cysteines was defined as fixed modification. Variable modifications were restricted to two per peptide. The precursor and fragment ion m/z ranges were limited from 350 to 1001 and 200 to 1700, respectively. Precursor charge states were set to 2-4 charges (Exploris 1-4 charges). An FDR threshold of 1% was applied for peptide and protein identifications. The mass accuracy for MS1 and MS2 were set to 15 ppm for timsTOF pro data and 12 ppm for Exploris data. Ion mobility windows were automatically adjusted by the software. The match between runs and RT-dependent cross-run normalization options were enabled. The software Perseus version 2.1.3.0^8^ was used for further data analysis. Enrichment analyses were performed with DAVID webtool using the gene ontology cellular component CC_direct and biological process BP_direct databases as well as the reactome database with default settings^9^. Bubble plots were created in python using the Matplotlib package^10^ . Bar and box plots were generated with Graphpad Prism version 10.6.1. Gene disease associations from DISGENET (https://www.disgenet.com) database were downloaded at 2026-01-20^11,12^.

References

1.	San Nicoló M, Mertzig S, Berghaus A, et al. Novel, standardized sample collection from the brain-nose interface. *Methods*. 2025;234:233-241. doi:10.1016/j.ymeth.2024.12.012

2.	Pigoni M, Wanngren J, Kuhn PH, et al. Seizure protein 6 and its homolog seizure 6-like protein are physiological substrates of BACE1 in neurons. *Mol Neurodegener*. 2016;11(1):67. doi:10.1186/s13024-016-0134-z

3.	Wiśniewski JR, Zougman A, Nagaraj N, Mann M. Universal sample preparation method for proteome analysis. *Nat Methods*. 2009;6(5):359-362. doi:10.1038/nmeth.1322

4.	Hughes CS, Moggridge S, Müller T, Sorensen PH, Morin GB, Krijgsveld J. Single-pot, solid-phase-enhanced sample preparation for proteomics experiments. *Nat Protoc*. 2019;14(1):68-85. doi:10.1038/s41596-018-0082-x

5.	Müller T, Kalxdorf M, Longuespée R, Kazdal DN, Stenzinger A, Krijgsveld J. Automated sample preparation with SP3 for low-input clinical proteomics. *Mol Syst Biol*. 2020;16(1):e9111. doi:10.15252/msb.20199111

6.	Cox J, Hein MY, Luber CA, Paron I, Nagaraj N, Mann M. Accurate proteome-wide label-free quantification by delayed normalization and maximal peptide ratio extraction, termed MaxLFQ. *Mol Cell Proteomics*. 2014;13(9):2513-2526. doi:10.1074/mcp.M113.031591

7.	Demichev V, Messner CB, Vernardis SI, Lilley KS, Ralser M. DIA-NN: neural networks and interference correction enable deep proteome coverage in high throughput. *Nat Methods*. 2020;17(1):41-44. doi:10.1038/s41592-019-0638-x

8.	Tyanova S, Temu T, Sinitcyn P, et al. The Perseus computational platform for comprehensive analysis of (prote)omics data. *Nat Methods*. 2016;13(9):731-740. doi:10.1038/nmeth.3901

9.	Sherman BT, Hao M, Qiu J, et al. DAVID: a web server for functional enrichment analysis and functional annotation of gene lists (2021 update). *Nucleic Acids Res*. 2022;50(W1):W216-W221. doi:10.1093/nar/gkac194

10.	Hunter JD. Matplotlib: A 2D Graphics Environment. *Comput Sci Eng*. 2007;9(3):90-95. doi:10.1109/MCSE.2007.55

11.	Piñero J, Corvi J, Rykova N, et al. DISGENET: Accelerating Data-Driven Discovery in Disease Genomics and Therapeutic Development. Preprint posted online January 5, 2026. doi:10.64898/2026.01.05.697749

12.	Piñero J, Ramírez-Anguita JM, Saüch-Pitarch J, et al. The DisGeNET knowledge platform for disease genomics: 2019 update. *Nucleic Acids Res*. 2020;48(D1):D845-D855. doi:10.1093/nar/gkz1021
